# Electronic Health Record Evaluation of the Preconception Medical Assessment for Maternal Morbidity

**DOI:** 10.64898/2026.03.02.26347437

**Authors:** Umair Khan, Saloni Shah, Gabriela Luna-Victoria, Lauren Groves, Diana Ramos, Marina Sirota, Tomiko T. Oskotsky

**Affiliations:** Bakar Computational Health Sciences Institute, University of California, San Francisco, San Francisco, CA, USA; Icahn School of Medicine at Mount Sinai, New York, NY, USA; Department of Obstetrics, Gynecology, and Reproductive Sciences, University of California, San Francisco, San Francisco, CA, USA; Office of the California Surgeon General, Sacramento, CA, USA; Department of Pediatrics, University of California, San Francisco, San Francisco, CA, USA; Division of Clinical Informatics and Digital Transformation, University of California, San Francisco, San Francisco, CA, USA

**Author notes:** Corresponding author:* Tomiko Oskotsky, MD, Bakar Computational Health Sciences Institute, University of California, San Francisco, 490 Illinois St, 2nd Floor, PO Box 2933, San Francisco, CA 94143. Co-lead authors (equal contribution) (Umair Khan, Saloni Shah). Co-senior authors (jointly supervised this work) (Diana Ramos, Marina Sirota, Tomiko T. Oskotsky).

## Abstract

**Importance:** Severe maternal morbidity is increasingly driven by chronic conditions present before pregnancy, but existing obstetric comorbidity indices rely on clinician documentation and are not designed for preconception use.

**Objective:** To determine whether conditions represented in a patient-facing preconception assessment are associated with severe maternal morbidity comparably with clinician-derived indices.

**Design:** Retrospective cohort study of deliveries from 1992 to 2025; scores were derived from diagnoses recorded at least 280 days before delivery.

**Setting:** Two deidentified electronic health record databases covering an academic and public health system in San Francisco and 5 other University of California medical centers.

**Participants:** Birthing individuals aged 12 to 55 years at delivery with at least 10 clinical events recorded before conception.

**Exposures:** Preconception Medical Assessment (PreMA) score (range, 0-8) and the Bateman (range, 0-51) and Leonard (range, 0-281) obstetric comorbidity indices, modeled per 1-SD increase.

**Main Outcomes and Measures:** Severe maternal morbidity and nontransfusion severe maternal morbidity within 7 days of delivery, defined by Centers for Disease Control and Prevention indicators.

**Results:** Among 58,549 deliveries (21,427 in the San Francisco cohort [mean (SD) maternal age, 33.4 (5.7) years]; 37,122 in the statewide cohort [33.1 (5.4) years]), each 1-SD increase in PreMA score was associated with severe maternal morbidity in the San Francisco cohort (relative risk [RR], 1.23; 95% CI, 1.19-1.28), similar to the Bateman (RR, 1.26; 95% CI, 1.22-1.29) and Leonard (RR, 1.26; 95% CI, 1.23-1.30) indices; statewide findings were consistent. Severe maternal morbidity rose from 6.6% at a score of 0 to 9.0% at 1 to 3 and 17.8% at 4 or greater (statewide, 3.8%, 5.3%, and 14.5%). Associations persisted across racial, ethnic, and neighborhood deprivation subgroups. Cardiovascular conditions (RR, 3.08; 95% CI, 2.50-3.80) and diabetes (RR, 1.83; 95% CI, 1.34-2.48) were most strongly associated with nontransfusion severe maternal morbidity.

**Conclusions and Relevance:** In this cohort study, higher scores on a patient-facing preconception assessment were associated with severe maternal morbidity, comparably with clinician-derived indices. Prospective studies are needed before scores are used clinically.

**Key Points:** *Question:* Is a brief patient-completed preconception assessment associated with severe maternal morbidity?

*Findings:* In this study of 58,549 deliveries across two California health systems, each standard deviation increase in the electronic health record-derived Preconception Medical Assessment score was significantly associated with severe maternal morbidity, with a magnitude similar to two established clinician-facing comorbidity indices. Severe maternal morbidity ranged from 6.6% among deliveries with a score of 0 to 17.8% among those scoring 4 or greater.

*Meaning:* Conditions captured by this patient-completed assessment may identify candidates for preconception counseling and chronic disease optimization.

## Introduction

Severe maternal morbidity (SMM) remains a major cause of maternal illness in the United States despite advances in obstetric care. SMM is associated with prolonged hospitalization, long-term health consequences, and substantial healthcare utilization. Many conditions that contribute to SMM — including chronic hypertension, diabetes, cardiovascular disease, and obesity — are present before pregnancy and may be amenable to evaluation or optimization before conception. Improving identification and management of these conditions before pregnancy therefore represents an important opportunity to reduce maternal complications.^1–6^

Existing obstetric comorbidity indices, such as the Bateman and Leonard indices, were designed primarily for specialist-based risk stratification and health services research.^7–9^ Clinically adapted indices and automated approaches applying natural language processing to clinician notes have since shown comparable performance in identifying patients at risk of SMM at admission for delivery.^10^ However, all of these tools depend on clinician documentation and medical record access, apply late in the care continuum, and are not intended as patient-facing tools for use before pregnancy.

To address this gap, the California Office of the Surgeon General developed the Preconception Medical Assessment (PreMA), a brief eight-item patient-completed questionnaire that presents established obstetric risk factors in clear, accessible language. Preliminary studies demonstrated that PreMA is feasible to administer, easy to understand, and acceptable to patients.^11^ By enabling individuals to recognize potential health risks before conception, PreMA aims to encourage earlier engagement with healthcare providers and facilitate timely counseling, referral, and optimization of chronic medical conditions before pregnancy. Whether this succinct patient-facing distillation of established obstetric risk factors retains sufficient clinical signal to identify SMM risk comparably to existing indices, however, has not been established.

In this multicenter retrospective cohort study, we evaluated the association between an electronic health record (EHR)–derived implementation of PreMA and SMM using two independent California health-system cohorts (eFigure 1). We compared these associations with those observed for the Bateman and Leonard obstetric comorbidity indices, and assessed consistency across racial, ethnic, and neighborhood deprivation subgroups. We hypothesized that higher preconception PreMA scores would be associated with increased SMM risk, with effect sizes comparable to the clinician-facing indices on which PreMA’s development was based.

## Methods

We conducted a retrospective observational study using two separate de-identified EHR databases. The UCSF-SFDPH database (hereafter, UCSF) combines EHR data from UCSF Health and the San Francisco Department of Public Health, running from 1988 and comprising over 6 million patients.^12^ The University of California Health Data Warehouse (hereafter, UC-wide), which aggregates medical records from over 8 million patients seen at six University of California medical centers (Davis, Irvine, Los Angeles, Riverside, San Diego, San Francisco), spanning 2012 to the present day, was also used.^13^ Both databases conform to the Observational Medical Outcomes Partnership (OMOP) Common Data Model (CDM) schema.^14^ All analysis of UCSF and UC-wide EHR data was performed under the approval of the Institutional Review Boards. All clinical data were de-identified and written informed consent was waived by the institutions. To avoid overlap, we excluded UCSF patients from our UC-wide queries (except where specified later).

The study population was identified using the birth admission codes from Alliance for Innovation on Maternal Health’s SMM Code List (v12-01-2022), which identify live births using International Classification of Diseases (ICD) codes, version 9 (ICD-9) and 10 (ICD-10).^15^ These codes were converted to standard OMOP concepts using the built-in concept relationship table and manually reviewed the results (eTable 1). Data were analyzed from 1992 to 2025. As specified by the inclusion criteria, mothers between the ages of 12 and 55 years at the time of delivery were selected for inclusion. To ensure adequate ascertainment of preconception health status, the cohort was restricted to pregnancies in which the birthing individual had at least 10 recorded clinical events (conditions, observations, or procedures in the OMOP schema) prior to conception. Because conception dates are not reliably recorded in EHRs, the preconception period was defined as more than 280 days before delivery. Gestational age is counted from the last menstrual period, which precedes conception by about 2 weeks, so this window ends before conception for nearly all deliveries and excludes conditions first diagnosed during pregnancy. Each delivery was treated as an independent observation. All eligible deliveries were included; no a priori sample size calculation was performed.

PreMA responses were operationalized by mapping questionnaire items to ICD-10 and Current Procedural Terminology (CPT) codes in consultation with the PreMA designers. Two published clinician-facing comorbidity indices (hereafter, Bateman^7^ and Leonard^8^), were used as baseline scores. All three indices were mapped to standard OMOP concepts using the concept relationship table and manually reviewed (eTable 2). For each delivery, preconception scores were calculated for PreMA (0 to 8), Bateman (0 to 51), and Leonard (0 to 281). To facilitate comparability across indices, each score was standardized within the analytic cohort and expressed as a z-score (mean 0, standard deviation 1).

The outcomes of interest were severe maternal morbidity (SMM) and non-transfusion SMM (NT-SMM). SMM was defined using the Centers for Disease Control and Prevention (CDC) indicators, a composite of life-threatening conditions and procedures occurring during the delivery hospitalization.^16^ SMM indicators occurring within seven days of the delivery date were identified. As before, we mapped indicators to standard OMOP concepts using the concept relationship table and manually reviewed the results (eTable 3). NT-SMM excluded SMM cases defined solely by blood transfusion.^17^

Demographic variables included maternal age at delivery, race, and ethnicity. Race and ethnicity were pulled from structured EHR fields using the categories available in each health system’s registration system. Race and ethnicity were examined because SMM is unequally distributed across racial and ethnic groups in the US, and because a screening instrument intended to expand access to preconception care should be evaluated for consistency across these groups. In our UC-wide analysis, socioeconomic context was assessed using the Area Deprivation Index^18^ (ADI), linked at the patient level and categorized into quintiles representing lowest (1-2), low (3-4), moderate (5-6), high (7-8), and highest (9-10) neighborhood deprivation. Since ADI was only available in the UC-wide dataset, we included UCSF patients in our UC-wide queries for this analysis. Conditions without a recorded diagnosis or procedure code were assumed to be absent. Missing race and ethnicity were retained as an unknown category, and deliveries without a linked Area Deprivation Index value were excluded from ADI-stratified analyses only. Because outcomes were ascertained within 7 days of delivery, no participants were lost to follow-up.

We estimated associations between each standardized risk score and maternal outcomes using regression models with the score entered as a continuous predictor. Logistic regression models were used to estimate odds ratios (ORs), consistent with prior studies of obstetric comorbidity indices.^7^ Poisson regression models with robust standard errors were used to estimate relative risks (RRs), facilitating direct comparison across outcomes and alignment with prior maternal health studies.^19^

Models were fit separately for each dataset (UCSF, UC-wide), score (PreMA, Bateman, Leonard) and outcome (SMM, NT-SMM). Primary models were unadjusted because the scores were evaluated as risk-stratification measures rather than causal exposures. Stratified analyses were conducted by race, ethnicity, and ADI quintile to assess heterogeneity across demographic and socioeconomic subgroups. To evaluate the contribution of individual PreMA questionnaire items, we fit Poisson regression models estimating the association between each item and SMM and NT-SMM. These models were adjusted for maternal age, race, and ethnicity. Maternal age was modeled using a spline term to account for its non-linear association with SMM and NT-SMM. Results were expressed as relative risks with 95% confidence intervals. Multiple testing correction was applied using the Benjamini-Hochberg false discovery rate procedure (q = 0.05).

To translate the score-outcome associations into clinically interpretable terms, we additionally summarized observed maternal outcomes across PreMA risk tiers. PreMA scores were grouped into three categories reflecting increasing preconception risk burden (0, 1 to 3, and 4 or greater), and the observed unadjusted proportions of SMM and NT-SMM were calculated within each tier separately in the UCSF and UC-wide cohorts. Two-sided 95% confidence intervals for each proportion were estimated using the Wilson score interval.

All analyses were conducted in Python version 3.10 (Python Software Foundation) using statsmodels version 0.14.4.^20^ All tests were 2-sided, and statistical significance was defined as a Benjamini-Hochberg false discovery rate–adjusted *P* < .05.

## Results

Table 1 summarizes the characteristics of the UCSF cohort (21,427 deliveries) and the UC-wide cohort excluding UCSF (37,122 deliveries), as well as the UC-wide cohort including UCSF for analyses requiring Area Deprivation Index data (n = 49,008). Maternal age distributions were similar across cohorts (mean [SD] age, 33.4 [5.7] years in UCSF data and 33.1 [5.4] years in UC-wide data), while the racial and ethnic composition differed modestly between UCSF and UC-wide (e.g., higher proportions of Asian patients in UCSF and higher proportions of White and Unknown race in UC-wide, Pearson’s chi-square P < .001).

**Table 1.** Patient characteristics in the UCSF and UC-wide EHR datasets. Baseline demographics for live births included in the analytic cohorts, shown for UCSF, UC-wide (excluding UCSF patients), and the UC-wide cohort including UCSF patients (used for Area Deprivation Index analyses). Reported P-values compare UCSF with the UC-wide cohort excluding UCSF patients.

|  |  | UCSF | UC-wide<br>(excluding<br>UCSF) | UC-wide<br>(including<br>UCSF) |
| --- | --- | --- | --- | --- |
| **n** |  | 21427 | 37122 | 49008 |
| **maternal age, mean (SD)**<br>[Welch's t-test, $P < .001$] | | 33.4 (5.7) | 33.1 (5.4) | 33.5 (5.4) |
| **race, n (%)**<br>[$\chi^2$, $P < .001$] | **American**<br>**Indian or**<br>**Alaska Native** | 105 (0.5) | 150 (0.4) | 174 (0.4) |
|  | **Asian** | 4092 (19.1) | 5283 (14.2) | 7847 (16.0) |
|  | **Black or**<br>**African**<br>**American** | 1511 (7.1) | 1823 (4.9) | 2389 (4.9) |
|  | **Multirace** | - | 2068 (5.6) | 2696 (5.5) |
|  | **Native**<br>**Hawaiian or**<br>**Other Pacific**<br>**Islander** | 439 (2.0) | 236 (0.6) | 298 (0.6) |
|  | **Other** | 4422 (20.6) | 5252 (14.1) | 6903 (14.1) |
|  | **Unknown** | 602 (2.8) | 2477 (6.7) | 2762 (5.6) |
|  | **White** | 10256 (47.9) | 19833 (53.4) | 25939 (52.9) |
| **ethnicity, n (%)**<br>[$\chi^2$, $P < .001$] | **Hispanic or**<br>**Latino** | 4800 (22.4) | 9464 (25.5) | 11250 (23.0) |
|  | **Not Hispanic or**<br>**Latino** | 16143 (75.3) | 26169 (70.5) | 35927 (73.3) |
|  | **Unknown** | 484 (2.3) | 1489 (4.0) | 1831 (3.7) |
| **UC location, n** | **1** | - | 4461 (12.0) | 4461 (9.1) |
| (%) | 2 |  | 9735 (26.2) | 9736 (19.9) |
|  | 3 |  | 7030 (18.9) | 7030 (14.3) |
|  | 4 |  | - | 11881 (24.2) |
|  | 5 |  | 15822 (42.6) | 15826 (32.3) |
|  | 6 |  | 74 (0.2) | 74 (0.2) |
| **Area Deprivation Index quintile, n (%)** | **lowest (1-2)** | - | 10022 (27.0) | 18792 (38.3) |
|  | **low (3-4)** |  | 9277 (25.0) | 10925 (22.3) |
|  | **moderate (5-6)** |  | 7836 (21.1) | 8310 (17.0) |
|  | **high (7-8)** |  | 5758 (15.5) | 6082 (12.4) |
|  | **highest (9-10)** |  | 2847 (7.7) | 3096 (6.3) |
|  | **(none)** |  | 1382 (3.7) | 1803 (3.7) |

Across both UCSF and UC-wide cohorts, higher comorbidity burden as measured by the Bateman index, Leonard index, and PreMA score was consistently significantly associated with increased risk of SMM and NT-SMM (Figure 1). In unadjusted logistic regression models, each standard deviation (SD) increase in score was associated with higher odds of both outcomes. Corresponding Poisson regression models yielded similar relative risk estimates. For SMM, per-SD increases in the Bateman, Leonard, and PreMA scores were associated with RR 1.26, 95% CI 1.22-1.29, *P* < .001; RR 1.26, 95% CI 1.23-1.30, *P* < .001; and RR 1.23, 95% CI 1.19-1.28, *P* < .001, respectively, in the UCSF data. Associations were comparable in magnitude for NT-SMM, with point estimates often slightly higher than those observed for SMM. Across all comparisons, the PreMA score demonstrated effect sizes similar to the established clinician-facing indices. Patterns were highly consistent between the UCSF and UC-wide analyses (Figure 1).

**Figure 1.**
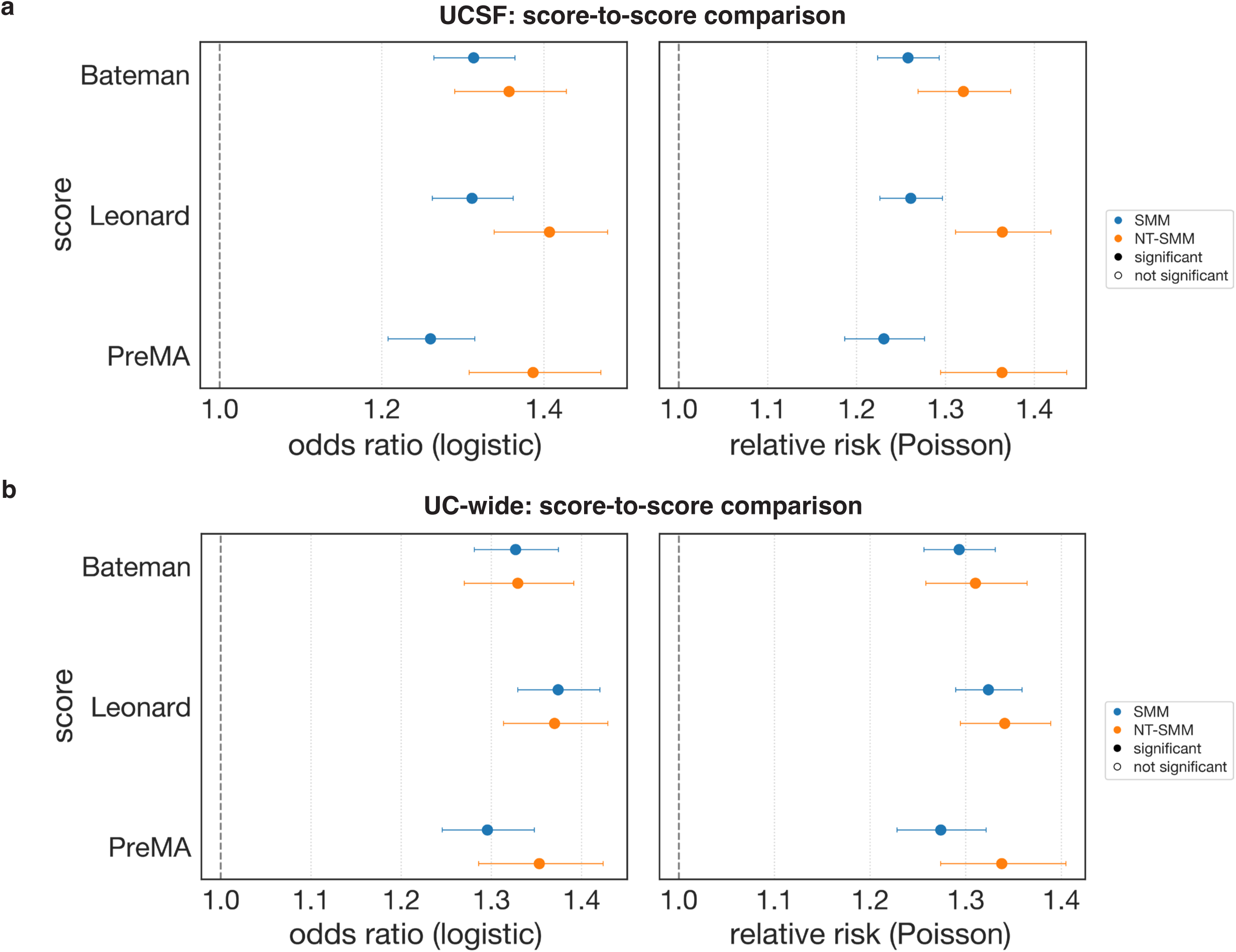
Score-to-score comparison of PreMA versus clinician-facing indices in UCSF and UC-wide cohorts. Forest plots showing associations per 1–SD increase in standardized Bateman, Leonard, and PreMA scores with SMM and NT-SMM in (a) UCSF and (b) UC-wide cohorts, using logistic regression (odds ratios) and modified Poisson regression (relative risks). Points represent effect estimates with 95% confidence intervals; marker fill indicates statistical significance as displayed in the figure legend.

When stratified by race, all three scores remained significantly positively associated with SMM and NT-SMM across most racial groups (Figure 2a). Effect estimates tended to be larger among American Indian or Alaska Native (UCSF PreMA NT-SMM RR 2.03, 95% CI 1.42-2.89, *P* < .001; UC-wide models did not converge), Black or African American (UCSF PreMA NT-SMM RR 1.33, 95% CI 1.19-1.49, *P* < .001; UC-wide PreMA NT-SMM RR 1.45, 95% CI 1.30-1.62, *P* < .001), and Native Hawaiian or Other Pacific Islander patients (UCSF PreMA NT-SMM RR 1.73, 95% CI 1.09-2.76, *P* = .023; UC-wide results not statistically significant) compared with White patients (UCSF PreMA NT-SMM RR 1.32, 95% CI 1.21-1.43, *P* < .001; UC-wide PreMA NT-SMM RR 1.29, 95% CI 1.20-1.39, *P* < .001), although confidence intervals were wider for smaller subgroups. Stratified analyses by ethnicity similarly demonstrated consistent associations for both Hispanic or Latino (UCSF PreMA NT-SMM RR 1.31, 95% CI 1.17-1.46, *P* < .001; UC-wide PreMA NT-SMM RR 1.46, 95% CI 1.34-1.59, *P* < .001) and non-Hispanic or Latino patients (UCSF PreMA NT-SMM RR 1.38, 95% CI 1.30-1.47, *P* < .001; UC-wide PreMA NT-SMM RR 1.29, 95% CI 1.22-1.37, *P* < .001) (Figure 2b). Across racial and ethnic strata, the PreMA score showed effect sizes comparable to or slightly exceeding those of the Bateman and Leonard indices, particularly for NT-SMM.

**Figure 2.**
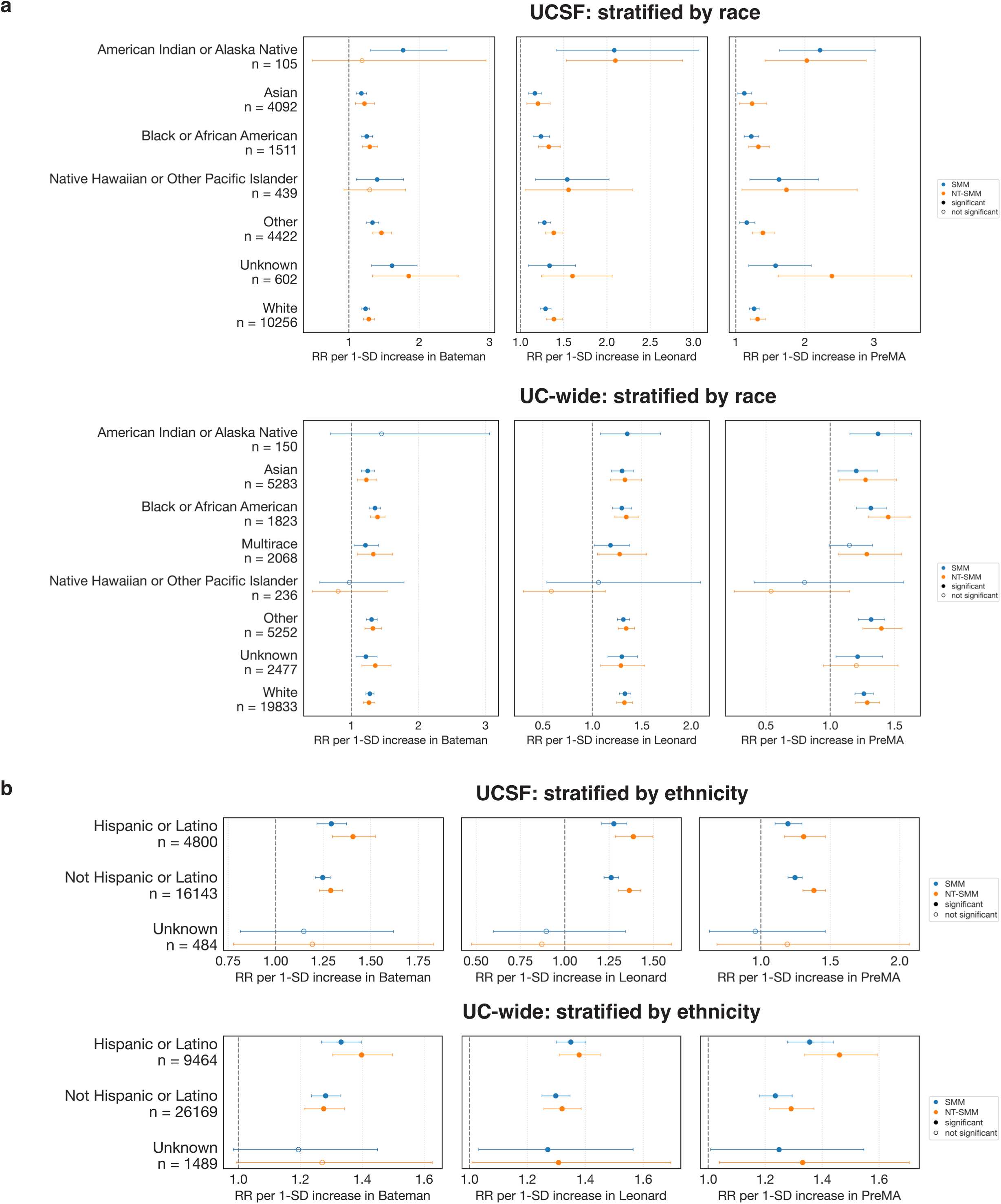
Stratified associations of comorbidity scores with severe maternal outcomes by race and ethnicity. (a) Relative risks for SMM and NT-SMM per 1-SD increase in Bateman, Leonard, and PreMA scores stratified by race within UCSF and UC-wide cohorts; (b) analogous stratified estimates by ethnicity. Effect estimates are shown with 95% confidence intervals.

Associations between comorbidity scores and outcomes were also examined across ADI quintiles in the UC-wide data (eFigure 2). For all three indices, higher scores were associated with increased risk of SMM and NT-SMM across levels of neighborhood deprivation. Effect estimates were generally larger in higher-deprivation quintiles, particularly for NT-SMM. The PreMA score was associated with an RR of 1.11, 95% CI 1.04-1.18, *P* = .001 in the lowest deprivation quintile and an RR of 1.31, 95% CI 1.19-1.43, *P* < .001 in the highest deprivation quintile for SMM. We noted that effect estimates decreased slightly for individuals in the highest ADI quintile compared to individuals in the high quintile.

To further characterize the contributions of individual PreMA questionnaire items, adjusted Poisson regression models were fit for each item, controlling for maternal age, race, and ethnicity (Figure 3). Several domains demonstrated strong associations with both outcomes, including cardiovascular conditions (UCSF NT-SMM RR 3.08, 95% CI 2.50-3.80, *P* < .001) and diabetes (UCSF NT-SMM RR 1.83, 95% CI 1.34-2.48, *P* < .001). Other domains, such as lung disease or breathing problems (UCSF NT-SMM RR 1.27, 95% CI 1.04-1.55, *P* = .032) and gastrointestinal disease or surgery (UCSF NT-SMM RR 1.17, 95% CI 0.99-1.39, *P* = .085), showed more modest associations, with some domains exhibiting weaker or non-significant associations after adjustment.

**Figure 3.**
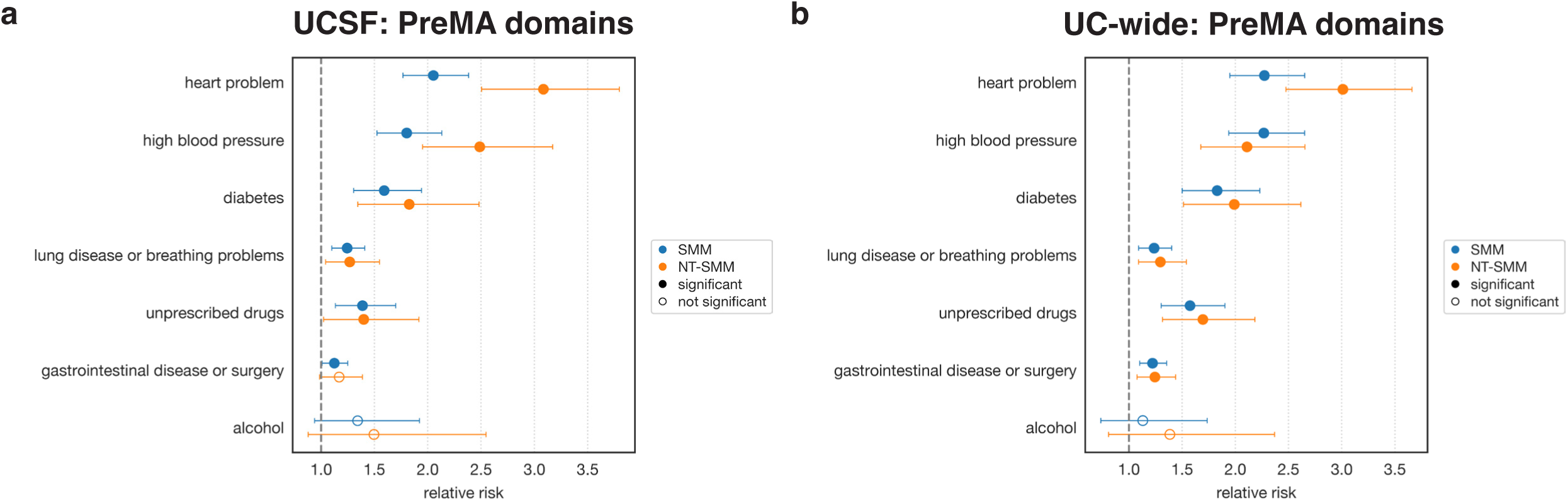
Analysis stratified by PreMA domains. PreMA domain-level associations with severe maternal outcomes in UCSF and UC-wide cohorts. Forest plots show adjusted relative risks for SMM (blue) and NT-SMM (orange) for each PreMA domain in (a) UCSF and (b) UC-wide. Estimates are displayed with 95% confidence intervals.

To express these associations in absolute terms, we summarized observed maternal outcomes across PreMA risk tiers (Table 2). In the UCSF cohort, observed SMM was 6.6% among deliveries with a PreMA score of 0, 9.0% at 1 to 3, and 17.8% at 4 or greater; the corresponding NT-SMM rates were 2.4%, 4.0%, and 8.5%. A consistent gradient was also present in the UC-wide cohort, where observed SMM rose from 3.8% at a score of 0 to 5.3% at 1 to 3 and 14.5% at 4 or greater, and NT-SMM increased from 1.9% to 2.8% to 9.5% across the same tiers. Absolute rates were higher overall in the UCSF cohort than in the UC-wide cohort across all tiers. Full event counts and 95% confidence intervals are provided in Table 2.

**Table 2.** Observed severe maternal morbidity by PreMA risk tier. Number of deliveries (n), outcome events, and observed rates of severe maternal morbidity (SMM) and non-transfusion severe maternal morbidity (NT-SMM) within PreMA score tiers (0, 1 to 3, and 4 or greater), shown separately for the UCSF and UC-wide cohorts. Percentages represent the unadjusted proportion of deliveries with each outcome within the tier, and 95% confidence intervals were calculated using the Wilson score method.

|  | **PreMA tier** | **n** | **% of cohort** | **SMM events** | **SMM % (95% CI)** | **NT-SMM events** | **NT-SMM % (95% CI)** |
| --- | --- | --- | --- | --- | --- | --- | --- |
| **UCSF** | 0 | 13,101 | 61.1 | 859 | 6.6<br>(6.2 – 7.0) | 309 | 2.4<br>(2.1 – 2.6) |
|  | 1-3 | 7,995 | 37.3 | 719 | 9.0<br>(8.4 – 9.6) | 318 | 4.0<br>(3.6 – 4.4) |
|  | 4+ | 331 | 1.5 | 59 | 17.8<br>(14.1 – 22.3) | 28 | 8.5<br>(5.9 – 12.0) |
| **UC-wide (excluding UCSF)** | 0 | 22,676 | 61.1 | 868 | 3.8<br>(3.6 – 4.1) | 428 | 1.9<br>(1.7 – 2.1) |
|  | 1-3 | 13,907 | 37.5 | 736 | 5.3<br>(4.9 – 5.7) | 393 | 2.8<br>(2.6 – 3.1) |
|  | 4+ | 539 | 1.5 | 78 | 14.5<br>(11.8 – 17.7) | 51 | 9.5<br>(7.3 – 12.2) |

All numerical model results are available in eTable 4.

## Discussion

In this multicenter retrospective cohort study, individuals with higher PreMA scores before pregnancy experienced significantly higher rates of severe maternal morbidity and non-transfusion severe maternal morbidity across two independent California health systems. These findings suggest that a brief patient-completed preconception assessment can identify individuals who may benefit from earlier counseling, optimization of chronic medical conditions, and referral before conception. Importantly, though PreMA was designed as a patient-completed screening tool rather than a clinician-facing risk index, it assessed risk of SMM comparably to two existing clinician-facing indices when using information available before pregnancy.

There is compelling evidence that preconception treatment and optimization of chronic conditions, including hypertension, diabetes, cardiovascular disease, yields substantial reductions in maternal morbidity.^21–23^ Thus, the preconception period represents an important opportunity to identify chronic medical conditions, help patients enter pregnancy in better health, and reduce their risk of maternal complications.^1,4–6^ The principal innovation of PreMA is not a new risk algorithm but the ability to extend validated obstetric risk assessment into the preconception period through a patient-completed instrument. Rather than improving statistical prediction, PreMA aims to move risk identification earlier in the care continuum, when counseling, optimization of chronic medical conditions, medication review, and pregnancy planning may still influence maternal outcomes.

Our findings demonstrate that the health conditions represented in PreMA are strongly associated with clinically meaningful maternal outcomes. Cardiovascular disease and diabetes emerged as particularly important contributors to SMM, reinforcing existing evidence that optimization of chronic medical conditions before pregnancy is an important component of maternal risk reduction. This is consistent with a national cohort analysis in which chronic hypertension and other hypertensive disorders of pregnancy were associated with SMM, particularly its cardiovascular forms.^24^ The increasing observed event rates across PreMA categories provide clinically interpretable evidence of a risk gradient, although prospective evidence is necessary before these categories are used for clinical triage.

These findings have several potential implications for clinical practice. Because PreMA is designed to be completed by patients before pregnancy, it may facilitate earlier recognition of modifiable risk factors among individuals who may not otherwise receive formal preconception counseling or who have not yet entered prenatal care. The questionnaire may also serve as an entry point into care for individuals seen in primary care, gynecology, family planning, community health, or other outpatient settings where pregnancy planning is discussed but formal obstetric risk assessment is not routinely performed. Earlier engagement is especially important for Black individuals and American Indian and Alaska Native individuals, who experience disproportionately high rates of severe maternal morbidity and maternal mortality.^1,4–6^ For example, a patient reporting chronic hypertension or diabetes could receive medication review, blood pressure or glycemic optimization, and referral to maternal-fetal medicine before conception. Importantly, PreMA is not intended to replace clinician judgment or existing risk assessment tools, but to promote patient engagement and timely referral into existing clinical pathways.

PreMA remained associated with SMM across racial, ethnic, and neighborhood deprivation subgroups, suggesting broad applicability. Future implementation studies should evaluate whether incorporating PreMA into community-based and primary care settings improves equitable access to preconception counseling.

Our per-item analyses suggest that PreMA may serve not only as a summary risk score but also as a framework for identifying specific domains that warrant targeted preconception counseling or intervention. Future work could explore weighted or adaptive versions of the instrument that emphasize the most prognostically informative components.

### Limitations

Several limitations should also be considered. First, this study retrospectively derived PreMA scores from EHR data rather than administering the questionnaire directly to patients.^25^ Consequently, the study evaluates the clinical relevance of the health conditions represented in PreMA but does not assess the usability, completion, or predictive performance of the patient-completed questionnaire in real-world settings. Second, although PreMA demonstrated associations with maternal outcomes similar to established clinician-derived obstetric comorbidity indices, similar effect sizes should not be interpreted as evidence of equivalent predictive performance because discrimination, calibration, and clinical utility were not evaluated. Third, although the study included diverse California populations, external validation in other healthcare systems and geographic regions will be important before widespread implementation. Fourth, requiring at least 10 clinical events before conception restricts the cohort to individuals with established health care use, who may differ from the populations PreMA is intended to reach. Code-based ascertainment likely undercounts preconception conditions and misses outcomes treated at outside facilities; such nondifferential misclassification would generally bias associations toward the null.

Future prospective studies should administer PreMA directly to patients before conception or early in pregnancy to evaluate questionnaire completion, discrimination, calibration, clinically meaningful thresholds, and integration into clinical workflows. Subsequent implementation studies should determine whether use of PreMA improves referral patterns, timely counseling, optimization of chronic medical conditions, and ultimately maternal outcomes.

## Conclusions

In this cohort study, scores on an EHR–derived implementation of PreMA were associated with SMM comparably to established clinician-facing indices, using only information available before conception. Prospective studies are needed to determine whether administering PreMA directly to patients improves preconception care and maternal outcomes.

## Supporting information

eTable 1

eTable 2

eTable 3

eTable 4

## Data Availability

The data that support the findings of this study are not openly available to individuals unaffiliated with UCSF due to the sensitivity of medical records, with the exception of collaborators. Individuals not affiliated with UCSF may set up an official collaboration with a UCSF-affiliated investigator by reaching out to the lead contact, Tomiko Oskotsky. UCSF-affiliated individuals may contact UCSF's Clinical and Translational Science Institute or the UCSF's Information Commons team for more information. UC-wide data is only available to UC researchers who have completed analyses in their respective UC first and have provided justification for scaling their analyses across UC health centers.

## Article Information

## Author Contributions

UK and SS contributed equally as co-first authors. DR, MS, and TTO contributed equally as co-senior authors. *Concept and design:* UK, DR, MS, TTO. *Acquisition, analysis, or interpretation of data:* UK, SS, GLV. *Manuscript drafting:* UK, SS, GLV. *Manuscript review:* all authors. *Statistical analysis:* UK. *Funding:* MS, TTO. *Administrative, technical, and material support:* LG, DR. *Supervision:* LG, DR, MS, TTO.

## Meeting Presentations

This work was presented in part at the 73rd Annual Scientific Meeting of the Society for Reproductive Investigation; March 24-28, 2026; San Juan, Puerto Rico; and has been accepted for oral presentation at the American Society for Reproductive Medicine Scientific Congress; October 26, 2026; Baltimore, Maryland.

## Data Sharing Statement

See Supplement 2.

## Additional Contributions

We acknowledge the use of the UCSF Information Commons computational research platform, developed and supported by the UCSF Bakar Computational Health Sciences Institute in collaboration with IT Academic Research Services, the Center for Intelligent Imaging Computational Core, and the CTSI Research Technology Program. We thank the Center for Data-driven Insights and Innovation at UC Health for technical support related to use of the UC Health Data Warehouse and the UC Data Discovery Platform.

## Additional Information

The authors used ChatGPT 5.2 (OpenAI; released December 11, 2025) and Claude Opus 4.6 (Anthropic; released February 5, 2026) to convert section outlines into initial drafts and to edit language and improve readability. These tools were not used to generate data, analyses, references, or figures. All authors reviewed, verified, and edited the resulting content and take full responsibility for the integrity of the manuscript.

