## Supplement 1 for "Electronic Health Record Evaluation of the Preconception Medical Assessment for Maternal Morbidity"

**Supplemental Material**

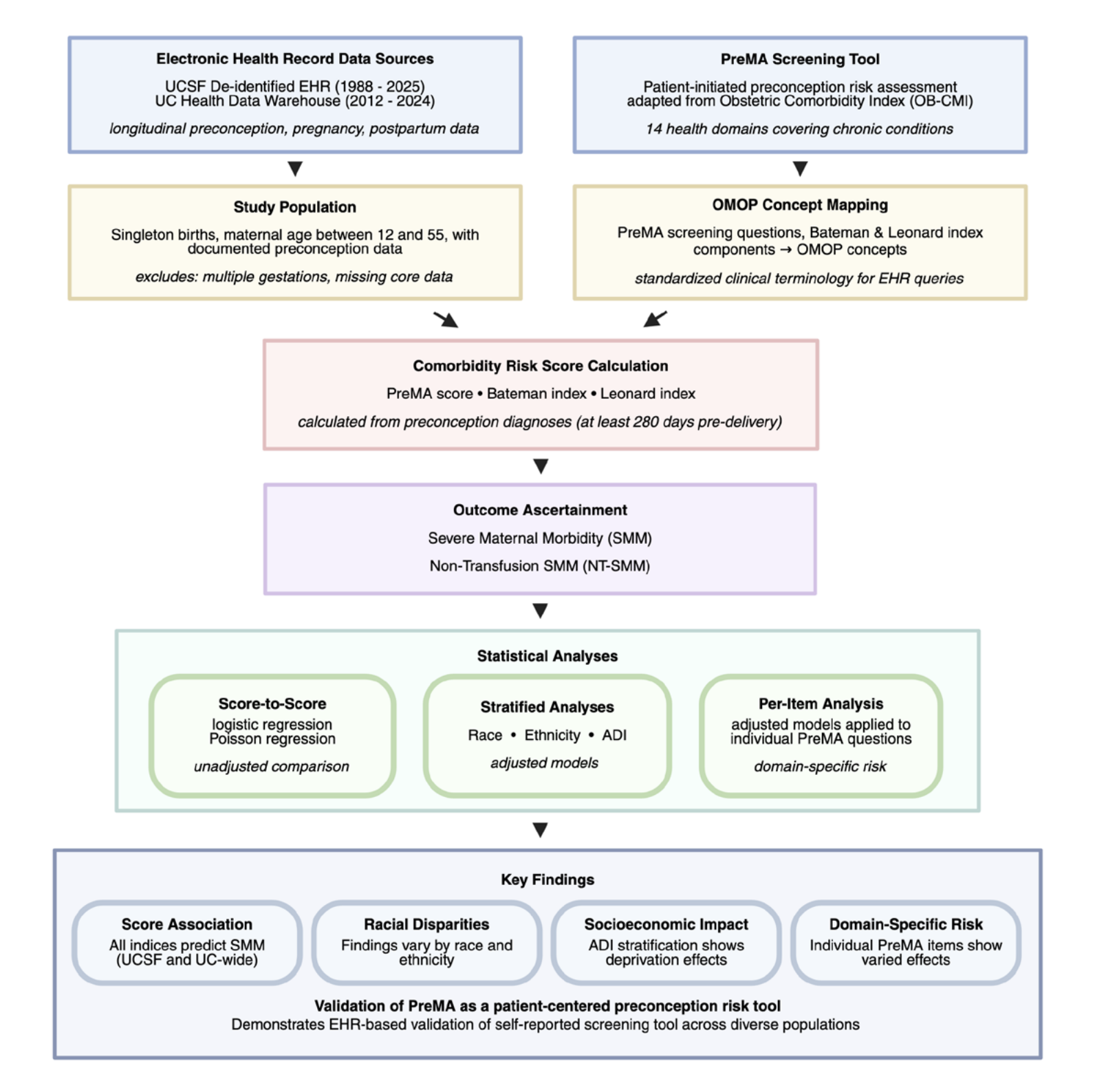

**eFigure 1. Study overview and analytic workflow.** Flow diagram depicting EHR data sources, study population definition, mapping of PreMA and comparator indices to OMOP concepts, preconception comorbidity score calculation, outcome ascertainment, and the three analytic components, culminating in key findings supporting EHR-based validation of PreMA.

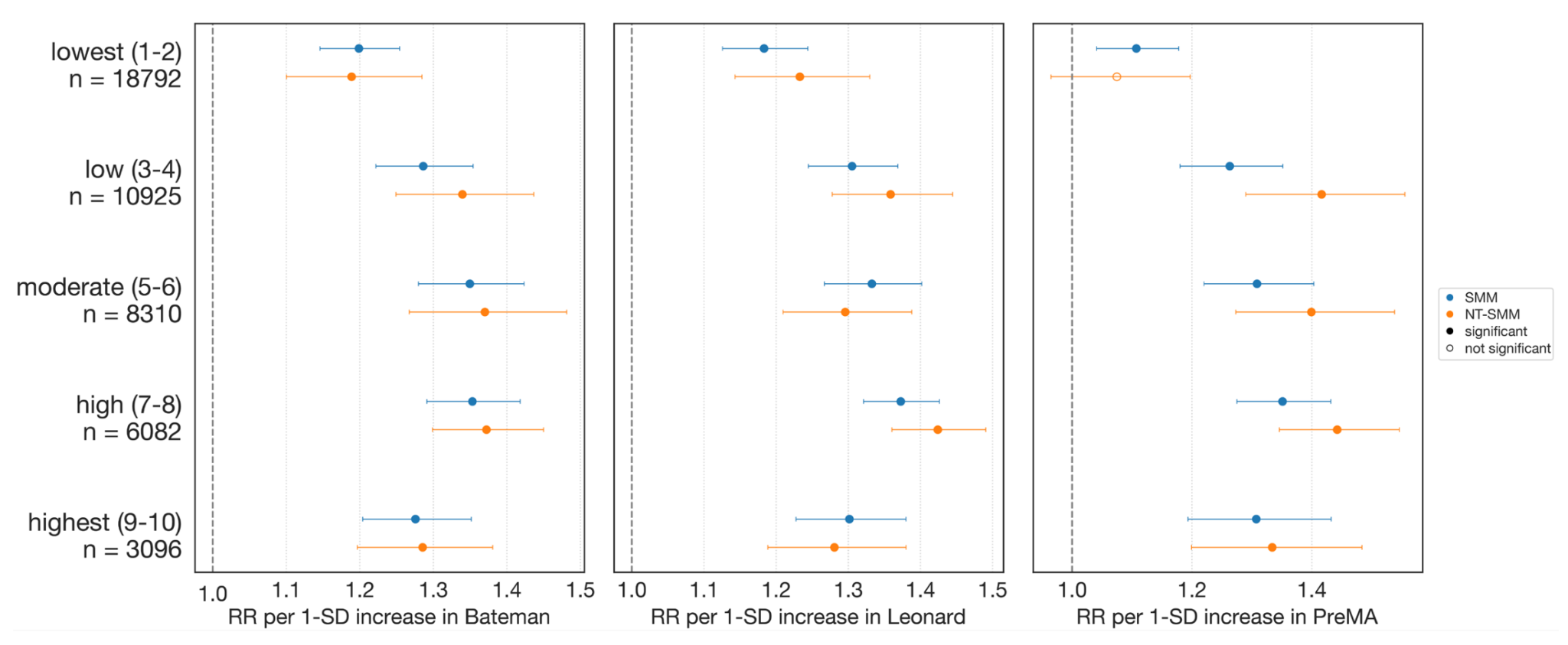

**eFigure2. Analysis stratified by Area Deprivation index.** Relative risks for SMM and NT-SMM per 1-SD increase in Bateman, Leonard, and PreMA scores stratified by Area Deprivation Index (ADI) quintile groupings (lowest to highest deprivation). Estimates are displayed with 95% confidence intervals.
