## Supplement 2 for "Electronic Health Record Evaluation of the Preconception Medical Assessment for Maternal Morbidity"

**Data Sharing Statement**

**Will individual participant data be available?** No.

**Data available:** Concept sets used to derive the PreMA, Bateman, and Leonard scores are available in the eTables. Analytic code is available upon request.

**Additional information:** The deidentified electronic health record data analyzed in this study were accessed under institutional data use agreements with the UCSF Information Commons and the University of California Health Data Warehouse and cannot be redistributed by the authors. Individuals not affiliated with UCSF may set up an official collaboration with a UCSF-affiliated investigator by reaching out to the lead contact. UCSF-affiliated individuals may contact UCSF’s Clinical and Translational Science Institute or UCSF’s Information Commons team for more information. UC-wide data are only available to UC researchers who have completed analyses in their respective UC first and have provided justification for scaling their analyses across UC health centers.
